# The Effect of Diabetes and Prediabetes on Anti-tuberculosis Treatment Outcomes: A Multicentric Prospective Cohort Study

**DOI:** 10.1101/2021.03.15.21253595

**Authors:** María B. Arriaga, Mariana Araújo-Pereira, Beatriz Barreto-Duarte, Betânia Nogueira, Maria Vitória C.N.S. Freire, Artur T.L. Queiroz, Moreno M.S. Rodrigues, Michael S. Rocha, Alexandra B. Souza, Renata Spener-Gomes, Anna Cristina C. Carvalho, Marina C. Figueiredo, Megan M. Turner, Betina Durovni, José R. Lapa-e-Silva, Afrânio L. Kritski, Solange Cavalcante, Valeria C. Rolla, Marcelo Cordeiro-Santos, Timothy R. Sterling, Bruno B. Andrade

## Abstract

**Background:** It is unclear whether diabetes or prediabetes drives adverse treatment outcomes and death in people with tuberculosis (PWTB).

**Methods:** Culture-confirmed PWTB, enrolled in the Regional Prospective Observational Research in Tuberculosis (RePORT)-Brazil cohort between 2015 and 2019 (n=756) were stratified based on glycemic status by baseline glycated hemoglobin levels. Unfavorable TB outcome was defined as treatment failure or modification, recurrence or death, whereas favorable outcome was cure or treatment completion. We validated the findings using data from PWTB reported to the Brazilian National System of Diseases Notification (SINAN) during 2015-2019 (n=20,989). Stepwise binary multivariable regression analysis models evaluated associations between glycemic status and unfavorable outcomes.

**Results:** In both cohorts, in univariate analysis, unfavorable outcomes were more frequently associated with drug resistance and HIV infection. Diabetes was associated with unfavorable outcomes in the RePORT (aOR: 2.85, p=0.001) and in SINAN (aOR: 1.56, p=0.040) cohorts. Furthermore, diabetes was associated with higher risk of death in both, RePORT-Brazil (aOR:3.23, p=0.006) and in the SINAN (aOR:2.75, p= 0.047) cohorts.

**Conclusion:** Diabetes was associated with an increased risk of unfavorable outcomes and mortality in Brazilian PWTB. Interventions to improve tuberculosis treatment outcomes in persons with diabetes are needed.

**40-word summary of the article’s main point:** In a multicenter prospective cohort study from Brazil, diabetes was associated with an increased risk of unfavorable treatment outcomes, including mortality, in pulmonary tuberculosis patients. These observations were validated in the Brazilian National Disease Notification System during the same period.

## INTRODUCTION

Diabetes (DM) is recognized by the World Health Organization (WHO) as a global epidemic [1]. This metabolic disease triples the risk of active tuberculosis (TB) in patients with latent *Mycobacterium tuberculosis* (Mtb) infection [2]. In 2019, approximately 400,000 people with tuberculosis (PWTB) worldwide were also diagnosed with diabetes [3]. Importantly, DM prevalence is increasing globally, including settings with a high (TB) burden, such as China and India [4].

The high prevalence of DM among TB patients (10% - 30%) in high TB burden countries and the negative impact of TB-DM comorbidity has been previously described by many groups [5-8] such as: higher mycobacterial loads and prevalence of sputum AFB positive, failure and death outcomes TB treatment, among others.

We have reported an association between DM and more severe TB clinical presentation (higher frequency of cough, night sweats, hemoptysis and malaise) [9], increased lung pathology reflected by severe radiographic manifestations (higher number of pulmonary lesions, including cavitation) [10], increased bacterial load in sputum [11] and delayed sputum conversion after anti-tuberculosis treatment initiation [12]. Furthermore, activation of tissue remodeling responses [12] and increased and persistent systemic inflammation [13] have been reported during treatment in TBDM patients. Thus, the increase in the number of people with DM may further complicate care and control of TB, especially in many areas with a high burden of both diseases [14].

There is also evidence that patients with concomitant TB and DM have an increased risk of unfavorable anti-tuberculosis treatment outcome such as failure, recurrence and death compared to normoglycemic patients [15-17]. However, the findings have not been consistent [18, 19]. In the present study we evaluated the effect of dysglycemia (DM or pre-DM) on anti-tuberculosis treatment outcomes in a prospective Brazilian cohort of patients with pulmonary TB (Regional Prospective Observational Research in Tuberculosis (RePORT)-Brazil), and also among PWTB reported to the Brazilian National TB Registry through the National System of Diseases Notification (SINAN).

## MATERIALS AND METHODS

### Ethics Statement

The study was conducted according to the principles in the Declaration of Helsinki. The RePORT-Brazil protocol was approved by the institutional review boards at each study site and at Vanderbilt University Medical Center. The protocol, informed consent, and study documents were approved by the institutional review boards at all study sites. The Institional Review Boards and the protocol approval numbers are as follows: (i) Instituto de Pesquisa Clínica Evandro Chagas, Fundação Oswaldo Cruz, Rio de Janeiro, Brazil (protocol no. 688.067), (ii) Secretaria Municipal de Saude do Rio de Janeiro, Brazil (protocol no. 740.554), (iii) Hospital Universitario Clementino Fraga Filho Rio de Janeiro, Brazil (protocol no. 852.519), (iv) Maternidade Climério de Oliveira, Universidade Federal da Bahia, Salvador, Brazil (protocol no. 723.168), (v) Fundação de Medicina Tropical Dr. Heitor Vieira Dourado, Manaus, Brazil (protocol no. 807.595). Participation was voluntary and written informed consent was obtained from all participants. Participation in RePORT-Brazil was voluntary, and written informed consent was obtained from all participants. All data extracted from SINAN were public and freely accessible. The anonymity of study subjects was preserved; all data were de-identified.

### Overall Study Design

RePORT-Brazil includes five study sites, located in cities with high TB burden: Salvador, Manaus, and Rio de Janeiro [20]. One main objective of the consortium is to describe the clinical outcomes of TB treatment and latent *Mtb* infection in Brazil. The details of the sites and representativeness of the RePORT-Brazil cohort to all TB patients in Brazil have been described previously [20].

For the current analysis, we included RePORT-Brazil participants with pulmonary TB ≥ 18 years old enrolled between June 2015 and June 2019 (**Figure1A**), with new or previously diagnosed culture-positive sputum (Lowenstein-Jensen medium or BD BACTEC MGIT), who received anti-tuberculosis treatment and had a treatment outcome recorded in the study database. Clinical and epidemiological information was collected at three in-person visits (i) anti-TB treatment initiation (baseline), (ii) two months after initiating treatment, and (iii) at the completion of anti-TB treatment. In addition, telephone follow-up was performed for all participants every 6 months until up to month 24 (when it corresponded). All data collected was stored in REDCap [21].

To establish the glycemic status of PWTB, baseline HbA1c in blood was measured. DM was defined according to American Diabetes Association (ADA) guidelines [22]. Patients were classified as having DM (HbA1c ≥6.5%), pre-diabetes (pre-DM; HbA1c1=5.7%-6.4%) or normoglycemia (HbA1c <5.7%). HbA1c≥5.7% was classified as dysglycemia.

SINAN is the Brazilian National System of Notification of Diseases [23], and includes diseases that require notification in all states and municipalities of Brazil; TB is a notifiable disease. The details of SINAN have been previously described [20]. PWTB reported to SINAN are diagnosed following the criteria in the Brazilian Manual of Recommendations for Tuberculosis control [24]. Diagnostic criteria include: (i) clinical and epidemiologic factors (presumptive diagnosis), (ii) bacteriology (sputum smear positive) or positive culture for Mtb (solid or liquid media), (iii) positive Xpert MTB RIF, (iv) chest radiography or (v) in the case of extrapulmonary TB, histopathology [24]. For each TB case reported, characteristics such as sex, age, race, education, alcohol consumption, illicit drug use, smoking habits, comorbidities, presence of HIV infection and test results, among others were also reported. Glycemic status was reported to SINAN as a diagnosis of diabetes (yes / no), not exclusively based on HbA1c level.

We used the SINAN information from the years 2015 to 2019 to match the enrollment period of RePORT-Brazil (see ref. [20] and **Figure 1B**). Of note, in 2014 the “Strategies for Care of people with Chronic diseases”, addressing comprehensive basic care for patients with TB and DM among others [25], was implemented in Brazil. In addition, we used the sub-set of data reported from Salvador, Manaus, and Rio de Janeiro, the cities with RePORT Brazil study sites.

**Figure 1:**
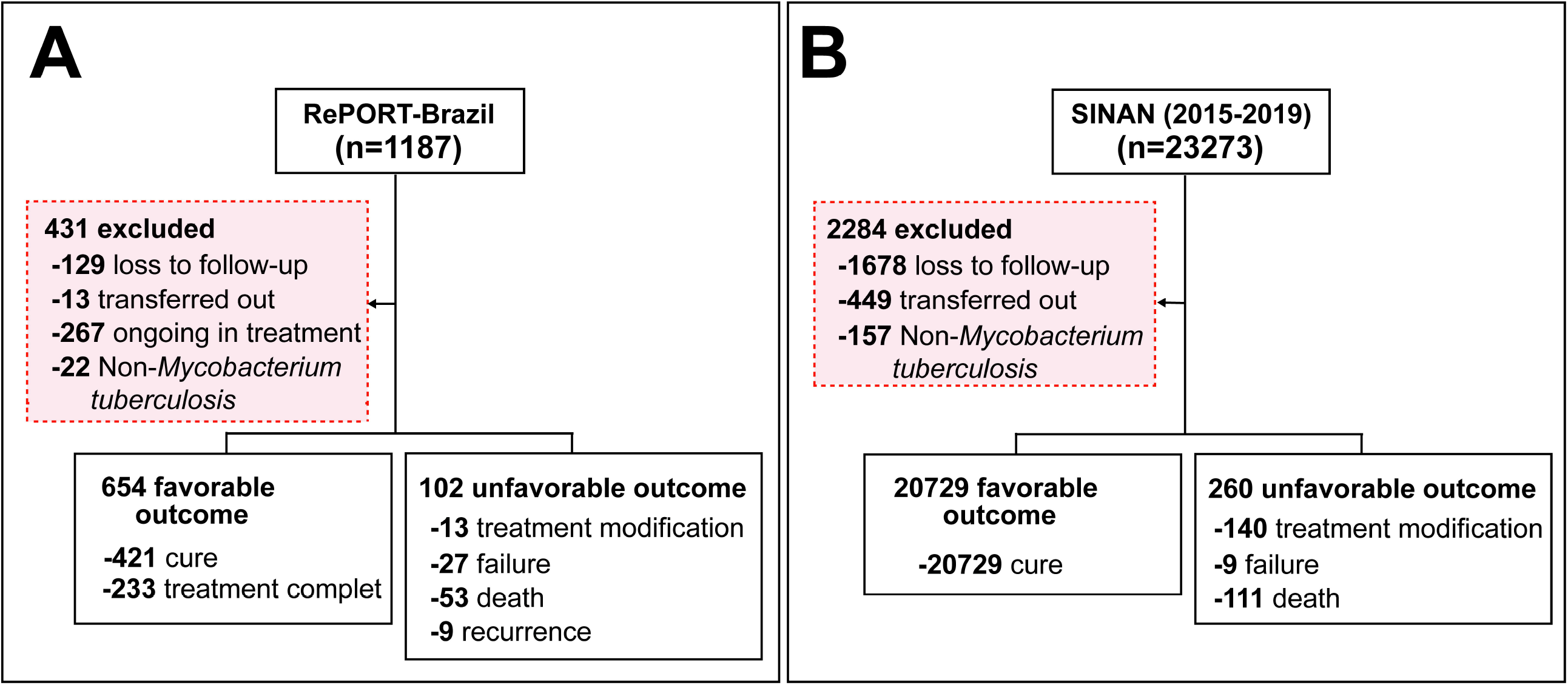
Study flowchart. Flowchart presenting the PWTB included and excluded from **(A)** RePORT-Brazil and PWTB reported to **(B)** SINAN between 2015-2019. Abbreviations: RePORT: Regional Prospective Observational Research for Tuberculosis. SINAN: Sistema de Informação de Agravos de Notificação. TB: Tuberculosis.

### Outcome definition

The primary outcome in this study was an unfavorable treatment outcome defined as treatment modification, treatment failure, recurrence, or death (during treatment). The secondary outcome was mortality (death for any reason during treatment). All definitions of outcome treatments were established in accordance with the Manual of Recommendations for the Control of TB of Brazil [24]. The treatment outcome definitions used in RePORT-Brazil and SINAN are depicted in **Supplementary Table 1**. A favorable outcome was defined as cure or treatment completion (defined as at least 90% of the total number of doses over one year for drug-susceptible TB and two years for TB-MDR). Patients who were lost to follow-up or transferred out or had a change in diagnosis (not a TB case) as the treatment outcome were excluded. The numbers of participants per outcome in this study are shown in more detail in **Figure 1A and 1B**.

**Table 1.** Characteristics of persons with TB in the RePORT-Brazil cohort by treatment outcomes.

| Characteristics | Unfavorable<br>(n=102) | Favorable<br>(n=654) | p-value |
| --- | --- | --- | --- |
| **Male, n (%)** | 71 (69.6) | 415 (63.5) | 0.267 |
| **Age, median (IQR)** | 38 (27-52) | 36 (25-49) | 0.208 |
| **Race, n (%)** |  |  | 0.050 |
| White | 18 (17.6) | 147 (22.5) |  |
| Black | 22 (21.6) | 168 (25.7) |  |
| Asian | 0 (0) | 4 (0.6) |  |
| **Pardo** | 57 (55.9) | 325 (49.8) |  |
| Indigenous | 5 (4.9) | 9 (1.4) |  |
| **Illiterate, n (%)** | 97 (95.1) | 626 (95.7) | 0.793 |
| **Smoking, n (%)<sup>a</sup>** | 66 (64.7) | 317 (48.5) | **0.030** |
| **Alcohol consumption, n (%)<sup>b</sup>** | 94 (92.2) | 537 (82.1) | **0.010** |
| **Illicit drug use, n (%)<sup>c</sup>** | 44 (43.1) | 176 (26.9) | **0.001** |
| **Prior TB, n (%)** | 17 (16.8) | 110 (16.9) | 1 |
| **Type of TB, n (%)<sup>d</sup>** |  |  | 0.235 |
| PTB | 87 (85.3) | 585 (89.4) |  |
| PTB+EPTB | 15 (14.7) | 69 (10.6) |  |
| **Abnormal X-ray, n (%)** | 95 (93.1) | 639 (97.7) | **0.020** |
| **Positive smear, n (%)** | 81 (79.4) | 528 (80.7) | 0.788 |
| **Resistance, n (%)** |  |  |  |
| Isoniazid | 13 (13.1) | 33 (5.2) | **0.006** |
| Rifampicin | 8 (8.1) | 11 (1.7) | **0.002** |
| Any drug<sup>e</sup> | 11 (11.1) | 71 (11.1) | 1 |
| Multidrug<sup>f</sup> | 6 (6.1) | 9 (1.4) | **0.009** |
| **DOT, n (%)** | 53 (51.9) | 368 (56.3) | 0.543 |
| **Glycemic status, n (%)** |  |  |  |
| Diabetes | 37 (36.3) | 128 (19.6) | **0.003** |
| Prediabetes | 25 (24.5) | 230 (35.2) | 0.43 |
| Normoglycemia | 40 (39.2) | 296 (45.3) | Ref. |
| **Dysglycemia, n (%)** | 62 (60.8) | 358 (54.7) | 0.284 |
| **Metformin use, n (%)<sup>g</sup>** | 2 (5.4) | 9 (7.0) | 0.516 |
| **Hemoglobin (g/dL), median (IQR)** | 10.6 (8.98-12.4) | 12.2 (10.95-13.4) | **<0.001** |
| **HbA1c (%), median (IQR)** | 6 (5.4-6.8) | 5.7 (5.4-6.2) | 0.054 |
| **HIV status, n (%)** | 45 (44.6) | 100 (15.5) | **<0.001** |
| **ART, n (%)<sup>h</sup>** | 13 (28.9) | 34 (33.7) | 0.702 |
| **Hypertension, n (%)** | 11 (10.8) | 56 (8.6) | 0.455 |
| **Other comorbidities, n (%)<sup>i</sup>** | 13 (12.7) | 32 (4.9) | **<0.001** |

**Table Note:** Data represent n. (%) or median with Interquartile range (IQR).
Favorable: cure/complete treatment. Unfavorable: failure, recurrence, treatment modification or death.
In RePORT-Brazil all patients had a positive culture at baseline.
<sup>a</sup>Smoking: Past or current cigarette smoker. <sup>b</sup>Alcohol consumption: Past or current any consumption of alcohol. <sup>c</sup>Illicit drug use: Past or current illicit drug use (marijuana, cocaine, heroin)
or crack). <sup>d</sup>All individuals from the RePORT cohort had a diagnosis of pulmonary tuberculosis, in some cases with presence in other anatomical sites. <sup>e</sup>Any drug (anti-tuberculosis) resistance except rifampicin and isoniazid: Pyrazinamide, ethambutol, streptomycin, kanamycin, ethionamide. <sup>f</sup>Multidrug: TB caused by Mycobacterium Tuberculosis (M. tuberculosis) strains that are resistant to at least both rifampicin and isoniazid. Only patients with the diabetes condition used metformin. <sup>g</sup>The frequency of use of metformin was calculated only in patients with diabetes. <sup>h</sup>ART frequency was calculated among the persons living with HIV. <sup>i</sup> It include cancer, kidney disease, chronic obstructive pulmonary disease, emphysema, and asthma. Abbreviations: TB: Tuberculosis. PTB: Pulmonary TB. EPTB: Extrapulmonary TB. HIV: Human Immunodeficiency Virus. DOT: Directly Observed Therapy. ART: Antiretroviral therapy.

### Data analysis

All analyses were pre-specified. Median and interquartile ranges (IQR) were used as measures of central tendency. Categorical variables were represented as percentages and compared using a two-sided Pearson’s chi-square test with Yates correction or the Fisher’s two-tailed test in 2×3 or 2×2 tables, respectively. Quantitative variables were compared using the Mann Whitney *U* test. We performed a logistic binary regression with backward stepwise selection in both the RePORT-Brazil and SINAN cohorts using variables with univariate p-value≤0.2 to identify independent associations between characteristics of TB patients (one model for each stratification strategy of the glycemic status: dysglycemia, diabetes, prediabetes, and HbA1c value) with the composite unfavorable treatment outcome or with mortality alone. Results from both regression approaches were presented in terms of point estimates and 95% confidence intervals (CI). P-values < 0.05 were considered statistically significant. Statistical analyses were performed using SPSS 25.0 (IBM statistics), Graphpad Prism 9.0 (GraphPad Software, San Diego, CA) and JMP 13.0 (SAS, Cary, NC, USA).

## RESULTS

### Factors associated with tuberculosis treatment outcomes in RePORT-Brazil

The RePORT cohort included 756 participants had culture-confirmed pulmonary TB who were treated with anti-tuberculosis drugs for at least 6 months. Patients were grouped according to treatment outcomes: favorable (n=654, 86.5%) and unfavorable (n=102, 13.5%) (**Figure1A**). The median age in RePORT cohort was 36 years [interquartile range (IQR): 25-49]. Most study participants were male in both groups. Patients with unfavorable outcomes more frequently reported current smoking (p=0.03), alcohol consumption (p=0.01) and illicit drug use (p=0.001) (**Table 1**). These patients also had higher frequency of resistance to isoniazid (p=0.006) or rifampicin (p=0.002), multi-drug resistance (p=0.009), diabetes (p=0.003) and HIV infection (p<0.001) (**Table 1**). The proportion of individuals with dysglycemia (i.e., with diabetes or prediabetes) did not differ significantly between those with unfavorable vs. favorable outcomes. In contrast, persons with unfavorable outcomes had lower hemoglobin levels (10.6 g/dL (IQR:8.98-12.4) vs. 12.2 g/dL (IQR:10.9-13.4) among those with favorable outcomes, p<0.001 (**Table 1**). In addition, no differences were found in the proportion of individuals with dysglycemia among those with drug-susceptible vs. multidrug resistant TB (**Supplementary Table 2)**.

**Table 2.** Characteristics of people with TB in the SINAN (2015-2019) cohort by outcomes.

| Characteristics | Unfavorable<br>(n=260) | Favorable<br>(n=23747) | p-value |
| --- | --- | --- | --- |
| **Male, n (%)** | 177 (68.1) | 13010 (62.8) | 0.082 |
| **Age, median (IQR)** | 39 (32-49) | 38 (28-52) | **0.217** |
| **Race, n (%)** |  |  | 0.674 |
| White | 64 (26.7) | 5116 (26.4) |  |
| Black | 56 (23.3) | 3495 (18.1) |  |
| Asian | 4 (1.7) | 170 (0.9) |  |
| Pardo | 116 (48.3) | 10500 (54.3) |  |
| Indigenous | 0 (0) | 62 (0.3) |  |
| **Illiterate, n (%)** | 35 (19.1) | 2422 (15.3) | 0.147 |
| **Smoking, n (%)<sup>a</sup>** | 68 (30) | 3540 (18) | **<0.001** |
| **Alcohol consumption, n (%)<sup>b</sup>** | 53 (23.3) | 2600 (13.1) | **<0.001** |
| **Illicit drug use, n (%)<sup>c</sup>** | 40 (17.8) | 1873 (9.6) | **<0.001** |
| **Prior TB, n (%)** | 101 (38.8) | 3209 (15.5) | **<0.001** |
| **Type of TB, n (%)<sup>d</sup>** |  |  | 0.107 |
| PTB | 247 (95) | 20071 (96.8) |  |
| EPTB | 4 (1.5) | 117 (0.6) |  |
| PTB+EPTB | 9 (3.5) | 541 (2.6) |  |
| **Abnormal X-ray, n (%)** | 220 (97.8) | 17914 (97.3) | 1 |
| **Positive smear, n (%)** | 157 (80.9) | 10718 (76.3) | 0.150 |
| **Positive culture, n (%)** | 116 (85.9) | 5877 (71.2) | **<0.001** |
| **Resistance, n (%)** |  |  | **<0.001** |
| Isoniazid | 34 (13.7) | 76 (0.3) | **<0.001** |
| Rifampicin | 7 (2.7) | 9 (0.03) | **<0.001** |
| Any drug<sup>e</sup> | 40 (15.4) | 145 (55.8) | **<0.001** |
| Multidrug<sup>f</sup> | 12 (4.6) | 13 (0.05) | **<0.001** |
| **DOT, n (%)** | 112 (64.7) | 10107 (60.6) | 0.275 |
| **Diabetes, n (%)** | 36 (15.4) | 2066 (10.4) | **0.015** |
| **HIV infection, n (%)** | 46 (21.3) | 1906 (10.6) | **<0.001** |
| **ART, n (%)<sup>g</sup>** | 23 (50) | 1225 (64.3) | **0.009** |
| **Hypertension, n (%)** | 5 (1.9) | 567 (2.7) | 0.564 |
| **Other comorbidities, n (%)<sup>h</sup>** | 23 (9.1) | 804 (4.0) | **<0.001** |

**Table Note:** Data represent n. (%) or median with Interquartile range (IQR).
Favorable: cure/complete treatment. Unfavorable: failure, recurrence, treatment modification or death.
<sup>a</sup>Smoking habits: Past or current cigarette smoker. <sup>b</sup>Alcohol consumption: Past or current any consumption of alcohol. <sup>c</sup>Illicit drug use: Past or current illicit drug use (marijuana, cocaine, heroin or crack). <sup>d</sup>Only individuals from the SINAN 2015-2019 cohort that had a diagnosis of pulmonary tuberculosis, in some cases with presence in other anatomical sites, were included. <sup>e</sup>Any drug (anti-tuberculosis) resistance except rifampicin and isoniazid: Pyrazinamide, ethambutol, streptomycin, kanamycin, ethionamide. <sup>f</sup>Multidrug: TB caused by *Mycobacterium Tuberculosis* strains that are resistant to at least both rifampicin and isoniazid. <sup>g</sup>ART frequency was calculated among the persons living with HIV. <sup>h</sup>It include cancer, kidney disease, chronic obstructive pulmonary disease, emphysema, and asthma.
Abbreviations: TB: Tuberculosis. PTB: Pulmonary TB. EPTB: Extrapulmonary TB. HIV: Human Immunodeficiency Virus. DOT: Directly Observed Therapy. ART: Antiretroviral therapy.

Regarding TB clinical presentation at study enrollment, patients who developed unfavorable outcomes more often were living with HIV (p<0.001) and presented with fatigue (p<0.001), whereas those who experienced a favorable treatment outcome more frequently presented with cough (p<0.001) and hemoptysis (p=0.043). (**Supplementary Figure 1A and B**).

We next evaluated the impact of baseline glycemic status on treatment outcomes. Dysglycemia was more frequent in TB patients who failed therapy (p=0.03) or died (p=0.043) during TB treatment (**Figure 2**). Baseline Hb1Ac values were distinguishable between the subgroups of participants who further developed favorable and unfavorable outcome when the outcomes were examined individually (treatment modification, failure, death, recurrence, and cure). Individuals with favorable treatment outcomes had lower HbA1c levels than those who experienced treatment modification (p=0.042), failure (p=0.024), and death (p=0.048) during treatment follow up. Among individuals with unfavorable outcomes, those who experienced treatment modification had lower Hb1Ac values than patients who had treatment failure (p=0.004) or died (p=0.016) (**Figure 2**). Furthermore, TB patients who experienced failure, recurrence or died exhibited a median HbA1c of 6g/dL (IQR:5.4-6.8).

**Figure 2.**
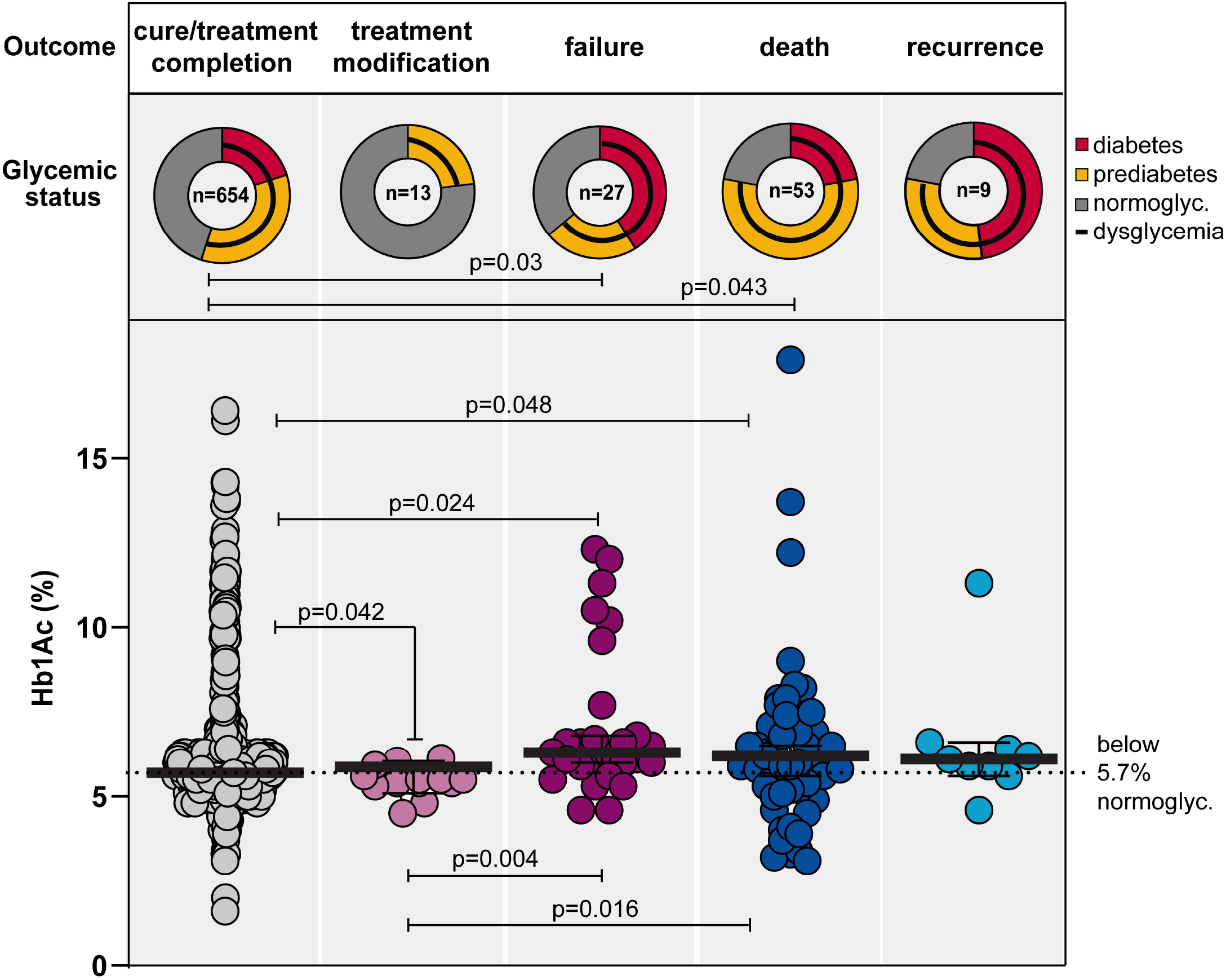
Distribution of the glycemic status and Hb1Ac levels according to treatment outcomes in TB patients in RePORT-Brazil. Frequency of PWTB with diabetes, prediabetes, normoglycemia and dysglycemia (diabetes + prediabetes) diagnosed using HbA1c levels is shown according to the TB treatment outcomes (cure, treatment modification, failure, death, and recurrence). Only comparisons (frequency dysglycemia status between TB treatment outcomes) with significant p-values are displayed. Scatter plots depicting the frequency of HbA1c values in TB patients according to TB treatment outcome. Lines represent median and interquartile range values. The differences in median values HbA1c between groups were compared using the Kruskal-Wallis test with Dunn’s multiple comparisons post-test. Only comparisons with significant p-values are displayed. Abbreviations: HbA1c: glycated hemoglobin, normoglyc.: normoglycemia

### Determinants of TB treatment outcomes in Brazilian National System of Notification of Diseases

In the SINAN cohort, with data also collected from 2015 to 2019, 20,989 PWTB were notified in the three cities where RePORT study sites are located. Males represented more than 60% of patients in both groups, and patients with unfavorable outcomes were older (45 years, IQR:32-58, p<0.001), more frequently reported smoking (p<0.001), alcohol consumption (p<0.001), illicit drugs use (p<0.001), and were less likely to report antiretroviral treatment (ART) (p=0.009) than those with favorable outcome. Unfavorable outcome was associated with previous TB (p<0.001), positive culture (p<0.001), diabetes (p=0.015), HIV co-infection (p<0.001), and other comorbidities (p<0.001) (**Table 2, Supplementary Figure 1C**).

Furthermore, we likewise evaluated the impact of glycemic status on the TB clinical presentation by the study participants in both cohorts (**Supplementary Figure 2>**). In RePORT patients, weight loss was more commonly observed in PWDM than in those with prediabetes or normoglycemia (p<0.001). In patients from SINAN, there was a higher proportion of PWTB with dysglycemia presenting with a positive smear at baseline than in those with normoglycemia (p<0.001). In addition, frequency of HIV infection was higher in the normoglycemic patients from the SINAN dataset (p <0.001). More details of other clinical factors are displayed in the **Supplementary Figure 2**.

### Multivariable logistic regression to assess association between glycemic status and tuberculosis treatment outcomes

A binomial logistic regression analysis was performed to test independent associations between the status of glycemia (dysglycemia, DM, pre-DM or HbA1c values) and treatment outcome in the RePORT cohort. DM was associated with unfavorable treatment outcomes independent of the other factors (adjusted OR [aOR]: 2.85, 95% CI: 1.57-5.17, p = 0.001) (**Figure 3, Model 2**). This analysis also was performed in SINAN cohort and DM was independently associated with unfavorable TB treatment outcomes (aOR: 1.56, 95% CI: 1.04-2.52, p = 0.04) (**Figure 3**).

**Figure 3.**
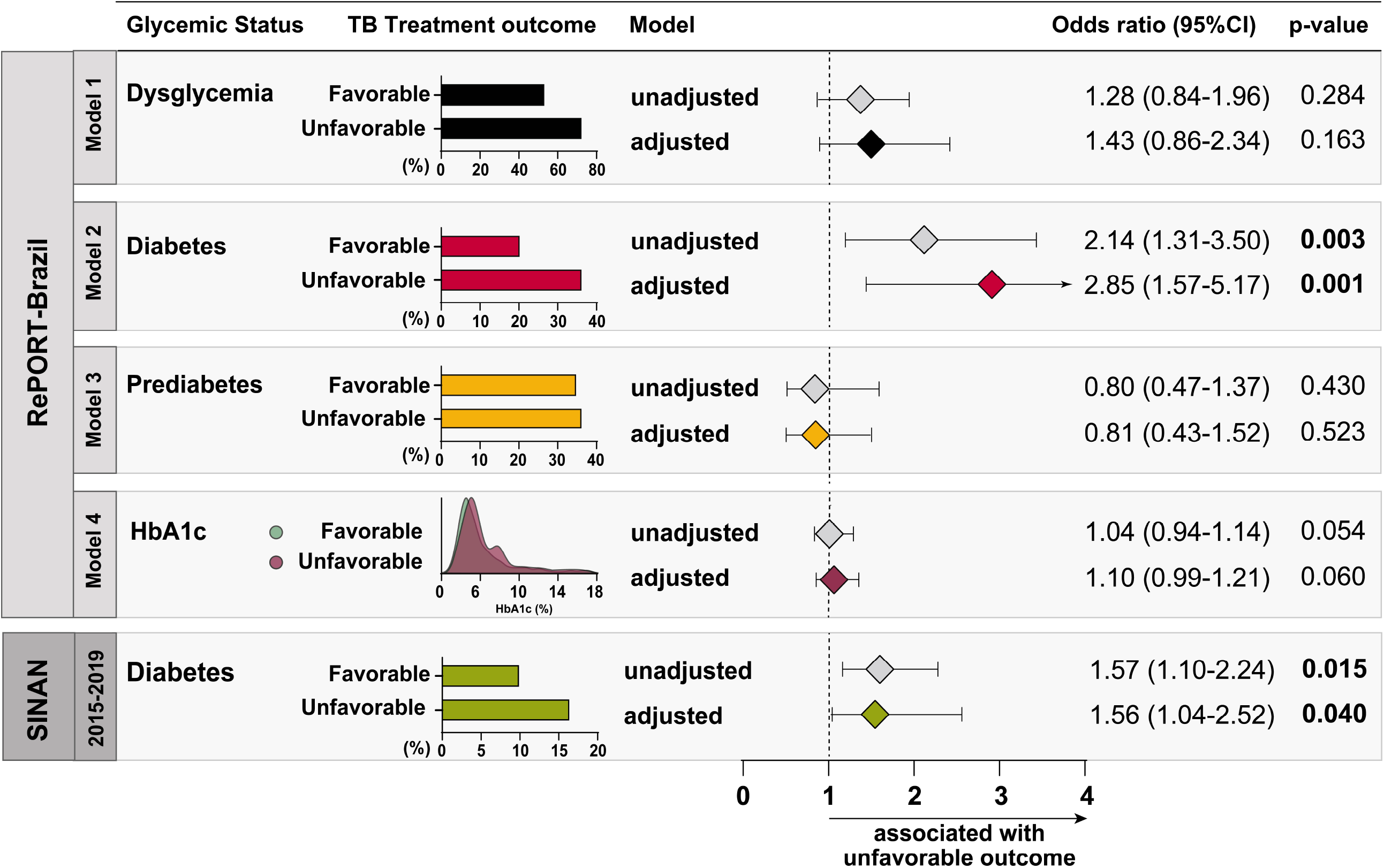
Association between glycemic status and TB treatment outcomes among TB patients from RePORT-Brazil and SINAN cohorts. In Report-Brazil cohort (upper panel) logistic regression models (backward stepwise regression) were performed to evaluate the independent associations between glycemic status of tuberculosis patients (Model 1: dysglycemia, Model 2: diabetes, Model 3: prediabetes and Model 4: increases of 1 unit in HbA1c level) and variables with p-value < 0.2 results in the univariate analyses (**Table 1**) and unfavorable treatment outcome (treatment modification, failure, recurrence, and death). In SINAN cohort (lower panel) logistic regression models (backward stepwise regression) were performed to evaluate the independent associations between diabetes of tuberculosis and variables with p-value<0.2 results in the univariate analyses (**Table 2**) and unfavorable treatment outcome (treatment modification, failure, and death). Abbreviations: RePORT: Regional Prospective Observational Research for Tuberculosis. SINAN: Sistema de Informação de Agravos de Notificação. TB: Tuberculosis. MDR-TB: multidrug-resistant tuberculosis. IDU: illicit drug use. PTB: pulmonary tuberculosis. HbA1c: glycated hemoglobin.

### Mortality during tuberculosis treatment

We next compared patients with favorable outcome versus those who died during TB treatment (n=55). Mortality was associated with male sex (p=0.039), increasing age (p=0.002), *pardo* race (61.8%, p=0.028), smoking (p=0.011) and illicit drug use (p=0.019), resistance to isoniazid (p=0.02), to rifampicin (p=0.004), multidrug resistance (p=0.049), HIV infection (p<0.001), diabetes (p=0.005) and with lower concentrations of hemoglobin (death: 9.45 g/dL, IQR:7.95-10.99; cure:12.2, IQR:11-13.4, p<0.001) and higher values of HbA1c (death:6.2%, IQR:5.2-7.1; cure: 5.7%, IQR: 5.4-6.2, p < 0.001) (**Supplementary Table 3**).

In the SINAN cohort, the subgroup of “death” as outcome showed the same statistically significant variables than those detected when we compared the composite unfavorable outcome *versus* cure (**Supplementary Table 4**). Patients who were successfully treated were younger (p<0.001), less likely to report smoking (p<0.001), alcohol consumption (p<0.002), illicit drug use (p<0.018), HIV infection (p<0.001) and other comorbidities (p=0.009) in comparison to those who died during TB treatment in both cohorts. In SINAN, patients who developed a favorable outcome had a lower frequency of diabetes (p=0.011) and higher frequency of ART use (p=0.001).

A logistic regression analysis was performed comparing cure and death and presented similar results to the first model exploring the composite unfavorable outcome. In the RePORT cohort, DM was once again strongly associated with mortality (aOR: 3.23, 95%CI: 1.39-7.43, p=0.006) (**Figure 4, Model 2**). In the SINAN cohort, DM was also significantly associated with death (aOR: 2.75, 95%CI: 1.02-7.42, p=0.047) (**Figure 4**).

**Figure 4.**
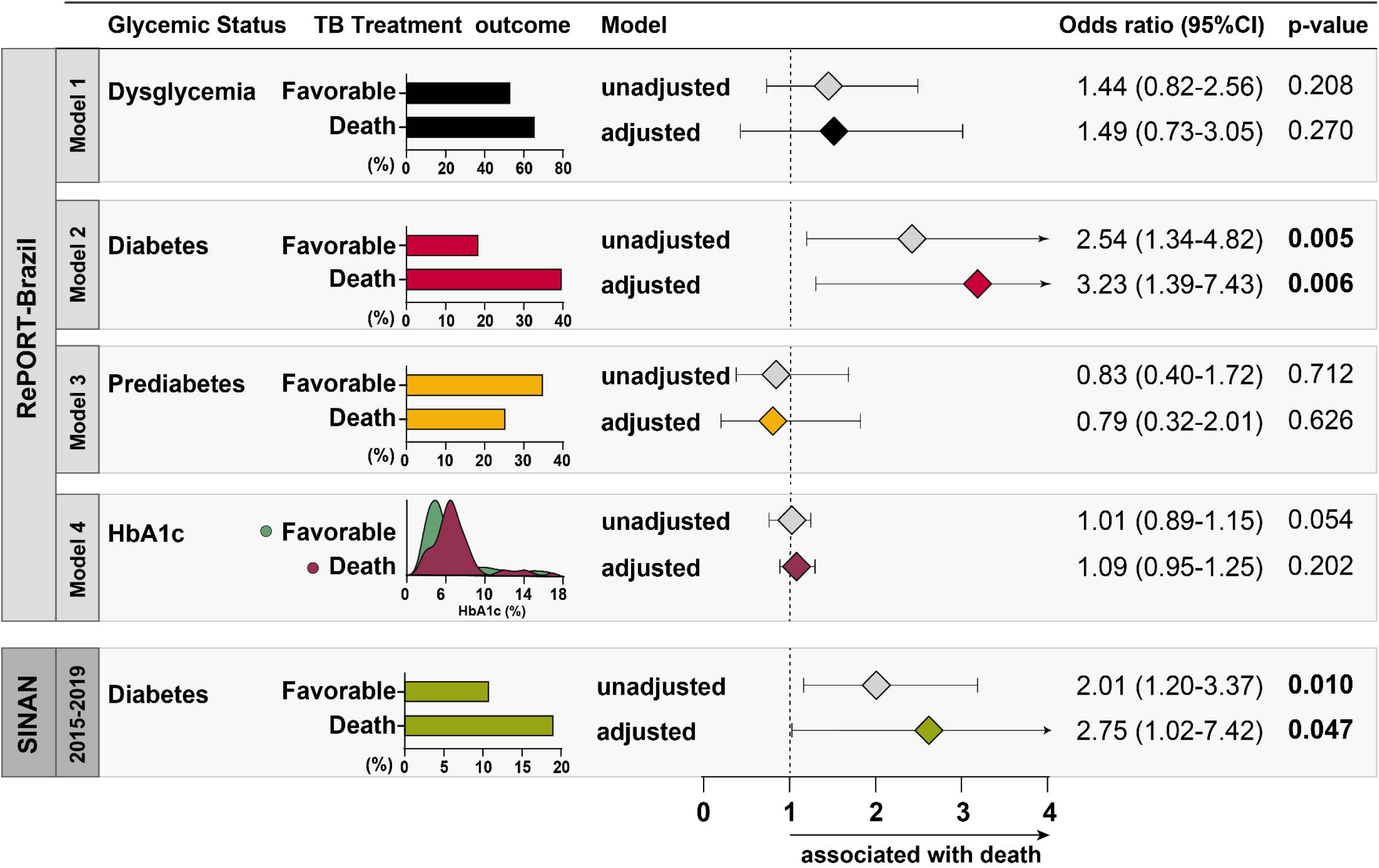
Association between glycemic status and death during anti-tuberculosis treatment among TB patients from RePORT-Brazil and SINAN cohorts. In Report-Brazil cohort (upper panel) logistic regression models (backward stepwise regression) were performed to evaluate the independent associations between glycemic status of tuberculosis patients (Model 1: dysglycemia, Model 2: diabetes, Model 3: prediabetes and Model 4: increases of 1 unit in HbA1c level) and variables with p-value < 0.2 results in the univariate analyses (**Supplementary Table 3**) and death. In SINAN cohort (lower panel) logistic regression models (backward stepwise regression) were performed to evaluate the independent associations between diabetes of tuberculosis patients in the period 2015-2019 and variables with p-value<0.2 results in the univariate analyses (**Supplementary Table 4**) and death. Abbreviations: RePORT: Regional Prospective Observational Research for Tuberculosis. SINAN: Sistema de Informação de Agravos de Notificação (Brazilian National System of Notification of Diseases) TB: Tuberculosis. HbA1c: glycated hemoglobin.

## DISCUSSION

There has recently been increased recognition of DM as an important risk factor for developing active TB and experiencing unfavorable TB treatment outcomes. [14-16, 26] As the prevalence of DM increases [27], particularly in developing countries, it is necessary to determine the public health impact of this syndemic in large populations. Our study analyzed data from PWTB in two data sources, a longitudinal cohort, and the nationwide disease notification system in the same period and the same three cities to determine the impact of DM on TB treatment outcomes in a Brazilian population.

It is interesting to note the factors that were consistently associated with poor outcomes in the two datasets analyzed, namely substance use (alcohol, tobacco, and illicit drug) and HIV infection, all in accordance with previous literature [19, 28] but different from other studies [29]. The analyses from RePORT also identified other drug resistance, anemia and normal chest radiograph as factors associated with worse outcomes. This latter observation may be due to the fact that RePORT performs systematic collection of these variables, which coincides with findings from other studies [19, 30].

In addition to the above factors, we found an association between higher HbA1c levels and treatment modification, treatment failure and death, when compared with the HbA1c levels among TB patients who were successfully treated, and this was reflected in the significantly higher proportion of PWD among those with unfavorable anti-tuberculosis treatment outcomes. In the RePORT cohort, the odds of having an adverse outcome was 2.85 times higher in DM patients compared to normoglycemic individuals. An odds ratio of 1.56 was found in the SINAN dataset. The relative lower detection of DM cases in SINAN dataset probably led to underestimation of the effect of DM in TB outcomes found in the analysis. Our results are similar to those from previous studies [13, 16]. The odds of death in the DM groups were 3.23 times higher than in normoglycemic patients in RePORT dataset and 2.5 in SINAN dataset. The odds ratio of death in the RePORT cohort was more compatible with what has been described in literature [16]. Previous studies showed how TBDM comorbidity is associated with a higher burden of immune pathology and systemic inflammation compared to TB normoglycemic patients [13, 31]. Furthermore, a defective regulation of the innate immune response in TBDM patients could maintain inflammatory foci despite anti-tuberculosis treatment, resulting in worse treatment results [13].

Moreover, all participants from RePORT had HbA1c measured at baseline visit, whereas in SINAN the diagnosis of DM was self-reported in most cases, possibly missing many diagnoses. Of note, a longitudinal cohort study done in Brazil found that 50% of individuals diagnosed with DM did not know about their diagnosis [32]. Thus, in the SINAN cohort, with increasing clinical evidence that diabetes is a risk factor for developing active TB [2] and that worsens the clinical TB presentation [9], screening for diabetes was increased over the recent years. But the problem of under-diagnosis and sub-notification still exists.

The findings presented here suggest that the impact of DM in TB in Brazil is underestimated. While the results reiterate the value of cohorts like RePORT in better delineating the local epidemiology, it highlights the importance of guidelines recommending laboratory investigation of DM in all TB incident cases and to establish specific treatment recommendations for this population. This implies long-term glycemic control that is essential to improve the outcome of TB treatment in patients with TBDM comorbidity [33]. The integrated care of individuals with both diseases has specific challenges such as the interaction between oral antidiabetic and anti-tuberculosis drugs [34] and the greater risk for adverse events [35].

We did not find an association between unfavorable outcome and death with pre-DM/dysglycemia in the RePORT cohort, but only with DM. Previous studies reported the normalization of glycemic levels during the treatment of TB, persons who were dysglycemic prior to anti-TB treatment [36, 37]. Therefore, the deleterious effects on the immune response caused by chronic DM hyperglycemia would not be observed in pre-DM, which could explain the lack of association with unfavorable and death outcomes [36, 37].

This study had several limitations. First, both RePORT and SINAN are observational cohorts; unmeasured or residual confounding could have explained the findings. Second, SINAN is a disease notification system that is not part of a study protocol. While SINAN represents TB cases from all of Brazil, there could be incomplete data collection or endpoint ascertainment, as well as different sources of information (i.e., self-reported *versus* information from medical charts) for different patients. Third, we did not have information on serum drug levels; low drug levels have been associated with unfavorable TB treatment outcomes, including in TB patients with advanced HIV and DM [38]. Nevertheless, we believe it brings valuable information on the impact of TB-DM on a population level.

Diabetes is a disease with growing prevalence and a major risk factor for unfavorable outcomes, including death during treatment, in individuals with TB, along with HIV infection and substance use. Actions prioritizing these groups are essential for the control of TB in Brazil.

## Data Availability

All data extracted from SINAN are public and freely accessible (http://www.portalsinan.saude.gov.br/dados-epidemiologicos-sinan). The data from RePORT Brazil can be available upon reasonable request to the corresponding author.

## NOTES

## Acknowledgments

The authors thank the study participants. We also thank the teams of clinical and laboratory platforms of RePORT Brazil. A special thanks to Elze Leite (FIOCRUZ, Salvador, Brazil), Eduardo Gama (FIOCRUZ, Rio de Janeiro, Brazil), Elcimar Junior (FMT-HVD, Manaus, Brazil), Hilary Vansell (VUMC, Nashville, USA) and Letícia C.M. Linhares (VUMC, Nashville, USA) for administrative and logistical support.

## Funding

The study was supported by the Intramural Research Program of the Fundação Oswaldo Cruz, Intramural Research Program of the Fundação José Silveira, Departamento de Ciência e Tecnologia (DECIT) - Secretaria de Ciência e Tecnologia (SCTIE) – Ministério da Saúde (MS), Brazil [25029.000507/2013-07] and the National Institutes of Allergy and Infectious Diseases [U01-AI069923]. M.B.A. received a fellowship from the Fundação de Amparo à Pesquisa da Bahia (FAPESB). M.A.-P. received a fellowship from Coordenação de Aperfeiçoamento de Pessoal de Nível Superior (Finance code: 001). B.B.A., J.R.L.S. and A.K. are senior investigators of the Conselho Nacional de Desenvolvimento Científico e Tecnológico (CNPq), Brazil.

## Disclaimer

The funder of the study had no role in the study design, data collection, or data analysis; however, a representative of the Brazilian Ministry of Health (A.K.) was involved in data interpretation and writing the report.

## Conflicts of interest

All authors: No reported conflicts of interest.

## Author contributions

Conceptualization, T.R.S., M.C.F., M.C.S., V.C.R., and B.B.A.; Data curation, M.B.A., M.A-P., .A.T.L.Q., M.M.S.R., and B.B.A.; Investigation, M.B.A., B.B.-D., M.A-P., B.N., A.B.S., M.S.R., A.B., RS-G., M.C.F., B.D., J.R.L.S., A.L.K., S.C., V.C.R., T.R.S., M.C.S., and B.B.A.; Formal analysis, M.B.A., M.A-P., A.T.L.Q., M.M.S.R. and B.B.A.; Funding acquisition, B.D., J.R.L.S., A.L.K., S.C., V.C.R., T.R.S., M.C.S., M.C.F., and B.B.A.; Methodology, M.B.A., B.B.-D., M.A-P and B.B.A.; Project administration, M.C.F., T.R.S., and B.B.A.; Resources, M.B.A., G.A., T.R.S., and B.B.A.; Software, M.B.A., M.A-P., A.T.L.Q., M.M.S.R., M.C.F., T.R.S., and B.B.A.; Supervision, T.R.S., and B.B.A.; Writing—original draft, M.B.A., B.B-D., M.A-P., B.N., M.V.C.N.S.F. and B.B.A.; Writing—review and editing, all authors. All authors have read and agreed to the submitted version of the manuscript.

## Notes

### Competing Interest Statement

The authors have declared no competing interest.

### Funding Statement

The study was supported by the Intramural Research Program of the Fundacao Oswaldo Cruz, Intramural Research Program of the Fundacao Jose Silveira, Departamento de Ciencia e Tecnologia (DECIT), Secretaria de Ciencia e Tecnologia (SCTIE), Ministerio da Saude (MS), Brazil [25029.000507/2013.07] and the National Institutes of Allergy and Infectious Diseases [U01AI069923]. M.B.A. received a fellowship from the Fundacao de Amparo a Pesquisa da Bahia (FAPESB). M.A.-P. received a fellowship from Coordenacao de Aperfeicoamento de Pessoal de Nivel Superior (Finance code: 001). B.B.A., J.R.L.S. and A.K. are senior investigators of the Conselho Nacional de Desenvolvimento Cientifico e Tecnologico (CNPq), Brazil.

### Author Declarations

The study was conducted according to the principles in the Declaration of Helsinki. The RePORT-Brazil protocol was approved by the institutional review boards at each study site and at Vanderbilt University Medical Center. The protocol, informed consent, and study documents were approved by the institutional review boards at all study sites. The Institional Review Boards and the protocol approval numbers are as follows: (i) Instituto de Pesquisa Clínica Evandro Chagas, Fundação Oswaldo Cruz, Rio de Janeiro, Brazil (protocol no. 688.067), (ii) Secretaria Municipal de Saude do Rio de Janeiro, Brazil (protocol no. 740.554), (iii) Hospital Universitario Clementino Fraga Filho Rio de Janeiro, Brazil (protocol no. 852.519), (iv) Maternidade Climério de Oliveira, Universidade Federal da Bahia, Salvador, Brazil (protocol no. 723.168), (v) Fundação de Medicina Tropical Dr. Heitor Vieira Dourado, Manaus, Brazil (protocol no. 807.595). Participation was voluntary and written informed consent was obtained from all participants. Participation in RePORT-Brazil was voluntary, and written informed consent was obtained from all participants. All data extracted from SINAN were public and freely accessible. The anonymity of study subjects was preserved; all data were de-identified.

