## Supplementary Material for "The Effect of Diabetes and Prediabetes on Anti-tuberculosis Treatment Outcomes: A Multicentric Prospective Cohort Study"

**Content:**

**Supplementary Table 1**

**Supplementary Table 2**

**Supplementary Table 3**

**Supplementary Table 4**

**Supplementary Figure 1**

**Supplementary Figure 2**

**Supplementary Table 1. Definition of tuberculosis treatment outcomes**

| Type of outcome treatment | Outcome tuberculosis treatment | RePORT | SINAN[a] |
|---|---|---|---|
| Favorable | **Cure** | Resolution of symptoms consistent with TB by the end of therapy. Patients without symptoms consistent with TB at the beginning of TB treatment could not have their clinical response evaluated. | Cure is established when pulmonary TB patients, initially sputum smear-positive, present, during treatment, with at least two negative sputum smears: one in the follow-up phase and the other at the end of treatment. |
| | **Completed treatment** | When the patient does not have treatment failure or loss to follow-up and has received at least 90% of the total number of doses of the standard recommended anti-TB therapy by the National TB Program for a period of up to one year for drug susceptible cases, and up to two years for MDR cases. For drug-susceptible TB, the drug regimen consisted of isoniazid, rifampicin, pyrazinamide, and ethambutol for 2 months, followed by isoniazid and rifampin for 4 months. For drug-resistant TB, treatment was based on drug susceptibility test results. | Completion of treatment based on clinical criteria and radiological: (i) when the patient has not undergone sputum examination due to absence sputum and are discharged based on clinical data and complementary exams, (ii) in cases of initially negative pulmonary tuberculosis; (iii) in cases of extrapulmonary tuberculosis |
| Unfavorable | **Treatment modification** | Switch to second line treatment[b] | When there is confirmation, through sensitivity testing or molecular rapid testing for tuberculosis, of resistance to any anti-tuberculosis medication, regardless of the treatment regimen treatment to be used, except in cases of monoresistance to ethambutol, pyrazinamide or streptomycin that keep using the basic regimen. |
| | **Failure** | Sputum smear- or culture-positive at month 5 or later during treatment. | When sputum positivity persists at the end treatment. Also classified as failure are patients who beginning of treatment are strongly positive (++ or +++) and maintain this until the fourth month, or those with initial positivity followed by negativity and new positivity for two consecutive months, from the fourth month of treatment. |
| | **Death** | A participant who dies for any reason after consenting to participate and prior to the end of the study. | On the occasion of knowledge of the patient's death during treatment. |
| | **Recurrence** | (i) Clinical or radiological evidence of active tuberculosis after completion of therapy (during the follow-up period) and met the definition for clinical cure, bacteriologic cure or | Relapse cases are not reported as treatment outcomes to SINAN. |

| | | | |
|---|---|---|---|
| | | bacteriologic status indeterminate with treatment complete. This includes persons who are AFB smear-positive or Xpert MTB/RIF-positive but AFB culture- negative. (ii) Positive culture for *M. tuberculosis* in liquid or solid medium (with any colony count), among patients who did not experience treatment failure and were considered "cured" or "bacteriologic status indeterminate with treatment completed", in a clinical sample collected after the end of the treatment, during the follow-up phase. | |
| | **Transferred out** | When the patient is transferred to another health service. | When the patient is transferred to another health service. |
| | **Loss to follow-up** | A participant who no longer participates in study visit follow-up or when an outcome cannot be assigned due to insufficient information. | A patient who has failed to attend the unit for more than 30 consecutive days after the expected return date. In treatment cases supervised, the period of 30 days is counted from the last date of taking the drug. |

**Table note:** [a]Described in the Manual of Recommendations for the Control of TB of Brazil. Abbreviations: RePORT: Regional Prospective Observational Research for Tuberculosis. SINAN: Sistema de Informação de Agravos de Notificação. MDR: Multidrug-Resistant. TB: Tuberculosis. [b]Described in the Treatment of Tuberculosis: Guidelines for National Programmes (The World Health Organization's Stop TB Department).

**Supplementary Table 2. Glycemic status of TB cases in RePORT and SINAN (2015-2019) cohorts by drug resistance TB**

| Glycemic status | Multidrug resistance | Drug-susceptible | p-value |
|---|---|---|---|
| **RePORT** | **n=15** | **n=721** | |
| Dysglycemia -n (%) | 9 (60.0) | 402 (55.8) | 0.799 |
| Diabetes -n (%) | 4 (26.7) | 156 (21.6) | 0.633 |
| Prediabetes -n (%) | 5 (33.3) | 246 (34.1) | 0.899 |
| Normoglycemia -n (%) | 6 (40.0) | 319 (44.2) | ref. |
| **SINAN** | **n=24** | **n=11159** | |
| Diabetes -n (%) | 14 (56.0) | 6000 (53.8) | 0.823 |
| Normoglycemia -n (%) | 11 (44.0) | 5159 (46.2) | ref. |

**Table note:** Data represent n. (%). Normoglycemia was used as a reference to calculates p-values of Dysglycemia, diabetes and prediabetes.

Abbreviations: ref.=reference

**Supplementary Table 3. Characteristics of patients in RePORT cohort according death and favorable treatment outcome**

| Characteristics | Death (n=55) | Favorable (n=674) | p-value |
|---|---|---|---|
| **Male, n (%)** | 43 (78.2) | 429 (63.6) | **0.039** |
| **Age, median (IQR)** | 40 (30-58) | 36 (25-49) | **0.002** |
| **Race, n (%)** | | | **0.028** |
| **White** | 10 (18.2) | 152 (22.6) | |
| **Black** | 8 (14.5) | 172 (25.6) | |
| **Asian** | 0 (0) | 4 (0.6) | |
| **Pardo** | 34 (61.8) | 336 (49.9) | |
| **Indigenous** | 3 (5.5) | 9 (1.3) | |
| **Iliterate, n (%)** | 52 (94.5) | 645 (95.7) | 0.727 |
| **Smoking, n (%)**[a] | 37 (67.3) | 328 (48.7) | **0.011** |
| **Alcohol consumption, n (%)**[b] | 48 (87.3) | 550 (81.6) | 0.363 |
| **Illicit drug use, n (%)**[c] | 23 (41.8) | 180 (26.7) | **0.019** |
| **Prior TB, n (%)** | 11 (20.4) | 117 (17.5) | 0.58 |
| **Type of TB, n (%)**[d] | | | 1 |
| **PTB** | 50 (90.9) | 604 (89.6) | |
| **PTB+EPTB** | 5 (9.1) | 70 (10.4) | |
| **Abnormal X-ray, n (%)** | 50 (90.9) | 658 (97.6) | 0.102 |
| **Positive smear, n (%)** | 45 (83.3) | 540 (80.2) | 0.722 |
| **Resistance, n (%)** | | | **<0.001** |
| **Isoniazid** | 7 (14) | 33 (5.1) | |
| **Rifampicin** | 5 (10) | 11 (1.7) | |
| **Any Drug**[e] | 6 (12) | 72 (11.2) | |
| **Multidrug**[f] | 3 (6) | 9 (1.4) | |
| **Glycemic status, n (%)** | | | |
| **Diabetes** | 22 (40) | 131 (19.6) | **0.005** |
| **Prediabetes** | 13 (23.6) | 235 (35.1) | 0.719 |
| **Normoglycemia** | 20 (36.4) | 303 (45.3) | Ref. |
| **Dysglycemia, n (%)** | 35 (63.6) | 366 (54.7) | 0.208 |
| **Hemoglobin (g/dL), median (IQR)** | 9.45 (7.95-10.99) | 12.2 (11-13.4) | **<0.001** |
| **HbA1c (%), median (IQR)** | 6.2 (5.2-7.1) | 5.7 (5.4-6.2) | **0.016** |
| **HIV status, n (%)** | 31 (56.4) | 102 (15.3) | **<0.001** |
| **ART, n (%)**[g] | 8 (25.8) | 35 (34) | 0.511 |
| **Hypertension, n (%)** | 8 (14.5) | 59 (8.8) | 0.149 |
| **Other comorbidities, n (%)**[h] | 9 (12.7) | 32 (4.9) | **0.012** |

**Table Note:** Data represent n. (%) or median with Interquartile range (IQR).
Favorable: cure/complete treatment.
[a]Smoking: Past or current cigarette smoker. [b]Alcohol consumption: Past or current any consumption of alcohol. [c]Illicit drug use: Past or current illicit drug use (marijuana, cocaine, heroin or crack). [d]All individuals from the RePORT cohort had a diagnosis of pulmonary tuberculosis, in some cases with presence in other anatomical sites. [e]Any drug (anti-TB) resistance except rifampicin and isoniazid: Pyrazinamide, ethambutol, streptomycin, kanamycin, ethionamide. [f].Multidrug: TB caused by *Mycobacterium Tuberculosis* strains that are resistant to at least both rifampicin and isoniazid. [g] ART frequency was calculated among the persons living with HIV. [h] It include cancer, kidney disease, chronic obstructive pulmonary disease, emphysema, and asthma.

Abbreviations: TB: Tuberculosis. PTB: Pulmonary TB. EPTB: Extrapulmonary TB. HIV: Human Immunodeficiency Virus. DOT: Directly Observed Therapy. ART: Antiretroviral therapy.

**Supplementary Table 4. Characteristics of patients in SINAN cohort according death and favorable treatment outcome**

| Characteristics | Death (n=111) | Favorable (n=23747) | p-value |
|---|---|---|---|
| **Male, n (%)** | 80 (72.1) | 13010 (62.8) | 0.050 |
| **Age, median (IQR)** | 48 (35-62) | 40 (28-54) | **<0.001** |
| **Race, n (%)** | | | 0.883 |
| **White** | 26 (26.0) | 5116 (26.4) | |
| **Black** | 19 (19.0) | 3495 (18.1) | |
| **Asian** | 2 (2.0) | 170 (0.9) | |
| **Pardo** | 53 (53.0) | 10500 (54.3) | |
| **Indigenous** | 0 (0.0) | 62 (0.3) | |
| **Illiterate, n (%)** | 15 (21.7) | 2422 (15.3) | 0.134 |
| **Smoking, n (%)**[a] | 28 (31.5) | 3540 (18.0) | <0.001 |
| **Alcohol consumption, n (%)**[b] | 22 (25.3) | 2600 (13.1) | **0.002** |
| **Illicit drug use, n (%)**[c] | 16 (17.8) | 1873 (9.6) | **0.018** |
| **Prior TB, n (%)** | 50 (45.0) | 3209 (15.5) | **<0.001** |
| **Type of TB, n (%)** | | | 0.185 |
| **PTB** | 105 (94.6) | 20071 (96.8) | |
| **EPTB** | 1 (0.9) | 117 (0.6) | |
| **PTB+EPTB** | 5 (4.5) | 541 (2.6) | |
| **Abnormal X-ray, n (%)** | 92 (97.9) | 17914 (97.3) | 1 |
| **Positive smear, n (%)** | 53 (68.8) | 10718 (76.3) | 0.136 |
| **Positive culture, n (%)** | 29 (85.3) | 5877 (71.2) | 0.086 |
| **Resistance, n (%)** | | | **<0.001** |
| **Isoniazid** | 24 (21.6) | 76 (0.3) | **<0.001** |
| **Rifampicin** | 3 (2.7) | 9 (0.03) | **<0.001** |
| **Any drug**[e] | 32 (28.8) | 145 (55.8) | **<0.001** |
| **Multidrug**[f] | 9 (8.1) | 13 (0.05) | **<0.001** |
| **DOT, n (%)** | 34 (54.8) | 10107 (60.6) | 0.365 |
| **Diabetes, n (%)** | 18 (18.9) | 2066 (10.4) | **0.011** |
| **HIV infection, n (%)** | 26 (31.3) | 1906 (10.6) | **<0.001** |
| **ART, n (%)**[g] | 14 (48.3) | 1225 (64.2) | **0.001** |
| **Hypertension, n (%)** | 4 (3.6) | 567 (2.7) | 0.550 |
| **Other comorbidities, n (%)**[h] | 10 (9.6) | 804 (4.0) | **0.009** |

**Table Note:** Data represent n. (%) or median with Interquartile range (IQR).
Favorable: cure treatment outcome

[a]Smoking: Past or current cigarette smoker. [b]Alcohol consumption: Past or current any consumption of alcohol. [c]Illicit drug use: Past or current illicit drug use (marijuana, cocaine, heroin, or crack). [e]ART frequency was calculated among the persons living with HIV. [e]include cancer, chronic obstructive pulmonary /emphysema, kidney disease, heart disease, liver disease, depression, dermatitis, and asthma. [f].Multidrug: TB caused by *Mycobacterium Tuberculosis* strains that are resistant to at least both rifampicin and isoniazid. [g] ART frequency was calculated among the persons living with HIV. [h] It include cancer, kidney disease, chronic obstructive pulmonary disease, emphysema, and asthma.

Abbreviations: TB: Tuberculosis. PTB: Pulmonary TB. EPTB: Extrapulmonary TB. HIV: Human Immunodeficiency Virus. DOT: Directly Observed Therapy. ART: Antiretroviral therapy.

### Supplementary Figures

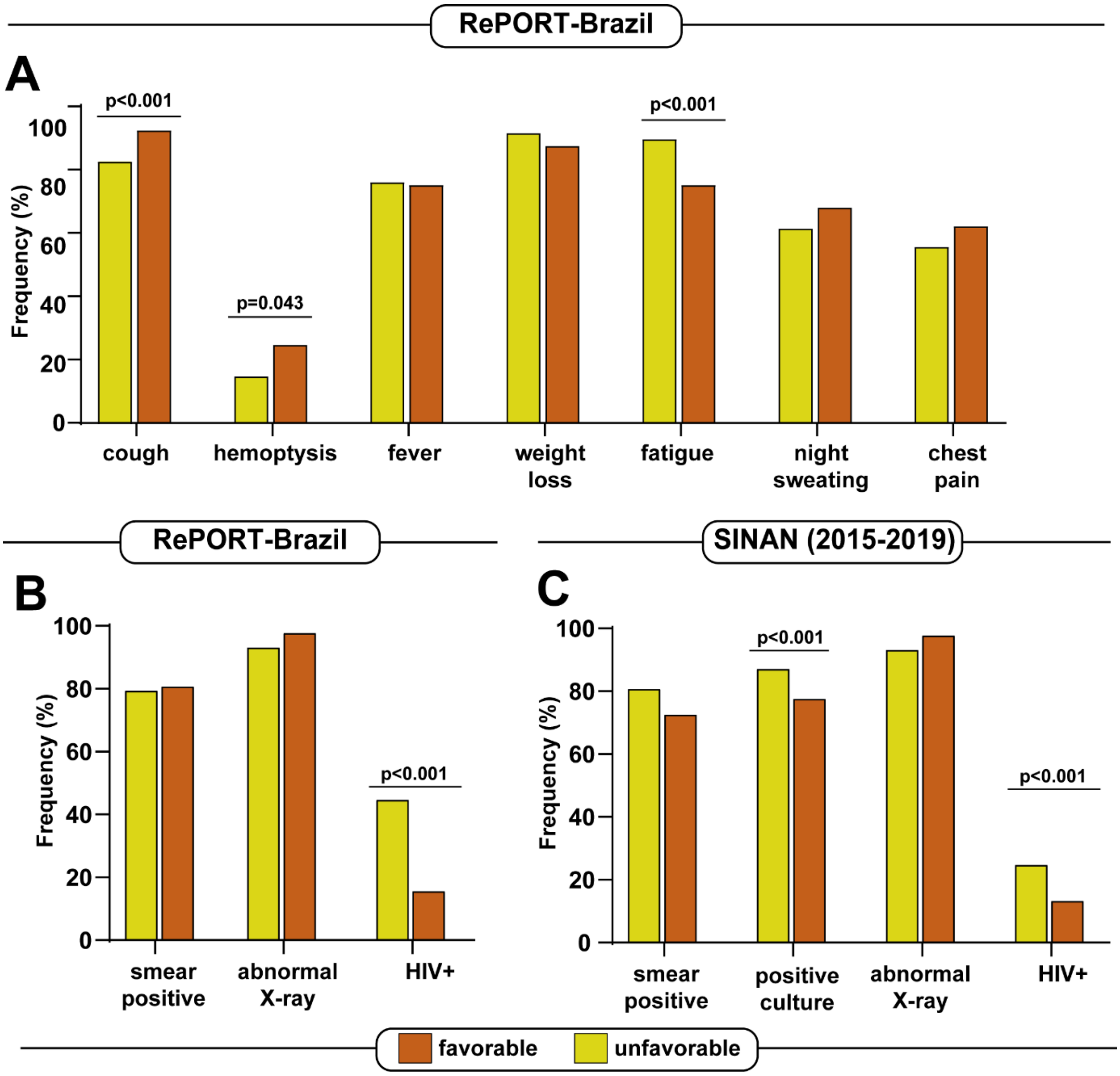


**Supplementary Figure 1. Frequency of clinical characteristics of TB patients from RePORT-Brazil and SINAN cohorts by treatment outcome**

**(A)**Frequency of symptoms, **(B)** smear positive, abnormal X -ray and HIV infection of TB cases reported cases according to the outcome in RePORT cohort. **(C)** frequency of smear positive, abnormal X -ray and HIV infection of TB cases according to the outcome reported to SINAN between 2010-2014. **(D)** frequency of smear positive, abnormal X -ray and HIV infection of TB cases according to the outcome reported to SINAN between 2015-2019. The data were compared between the groups using the Fisher's exact test (2×2 comparisons). Only comparisons with significant p-values are displayed.

Abbreviations: RePORT: Regional Prospective Observational Research for Tuberculosis. SINAN: Sistema de Informação de Agravos de Notificação (Brazilian National System of Notification of Diseases). TB: Tuberculosis.

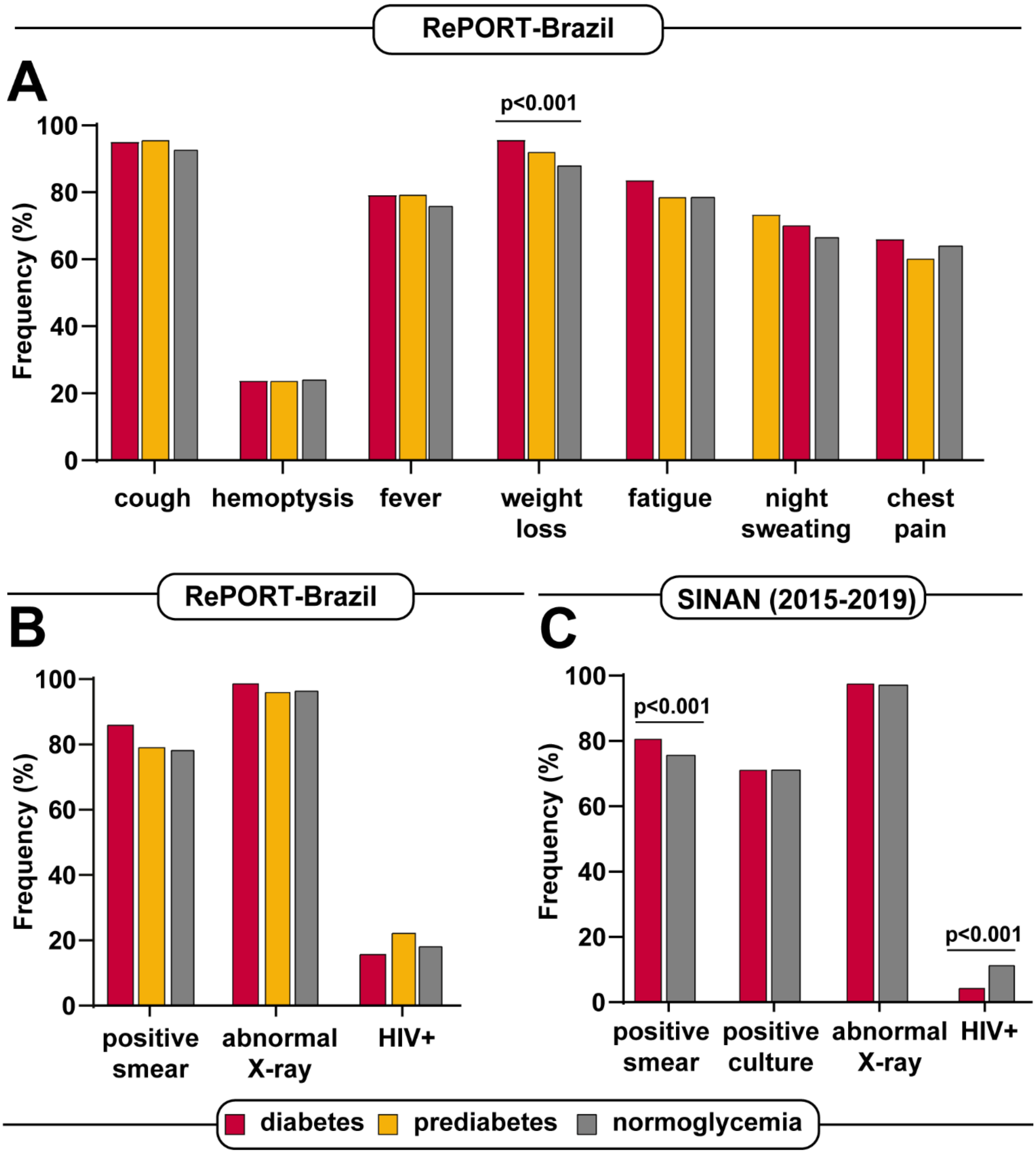


**Supplementary Figure 2**. **Frequency of clinical characteristics of TB patients from RePORT-Brazil and SINAN cohorts by glycemic status**

**(A)**Frequency of symptoms, **(B)** smear positive, abnormal X -ray and HIV infection of TB cases reported cases according to glycemic status (diabetes, prediabetes and normoglycemia) in RePORT cohort. **(C)** frequency of smear positive, abnormal X -ray and HIV infection of TB cases according to diabetes condition reported to SINAN between 2010-2014. **(D)** frequency of smear positive, abnormal X -ray and HIV infection of TB cases according to diabetes condition reported to SINAN between 2015-2019. The data were compared between the groups using Chi-square test with Yates correction or Fisher's two-tailed test in 2x3 or 2x2, respectively Only comparisons with significant p-values are displayed.

Abbreviations: RePORT: Regional Prospective Observational Research for Tuberculosis. SINAN: Sistema de Informação de Agravos de Notificação (Brazilian National System of Notification of Diseases). TB: Tuberculosis.
